# Phenotypic distinctions of *BLM-* and *RMI1-*associated Bloom syndrome

**DOI:** 10.1101/2021.11.02.21265560

**Authors:** Ipek Ilgin Gönenc, Nursel H. Elcioglu, Carolina Martinez Grijalva, Seda Aras, Nadine Großmann, Inka Praulich, Janine Altmüller, Silke Kaulfuß, Yun Li, Peter Nürnberg, Peter Burfeind, Gökhan Yigit, Bernd Wollnik

## Abstract

Bloom syndrome (BS) is an autosomal recessive disease with characteristic clinical features of primary microcephaly, growth deficiency, skin lesions, cancer predisposition, and immunodeficiency. Here, we report the clinical and molecular findings of eight patients from six families diagnosed with BS. We identified causative mutations in all families, three different homozygous mutations in *BLM* and one causative homozygous mutation in *RMI1*. The homozygous c.581_582delTT (p.Phe194*) and c.3164G>C (p.Cys1055Ser) mutations in *BLM* have already been reported in BS patients, while the c.572_573delGA (p.Arg191Lysfs*4) is novel. Interestingly, whole-exome sequencing revealed a homozygous loss-of-function mutation in *RMI1* in two BS patients of a consanguineous Turkish family. All BS patients had primary microcephaly, intrauterine growth delay, and short stature, presenting the phenotypic hallmarks of BS. However, a narrow face, skin lesions, and upper airway infections were observed only in some of the patients. Overall, patients with homozygous *BLM* mutations had a more severe BS phenotype compared to patients carrying the homozygous *RMI1* mutation, especially in terms of immunodeficiency and associated recurrent infections. Low-level immunoglobulins were observed in all *BLM*-mutated patients, emphasizing the immunodeficiency profile of the disease, which should be considered as an important phenotypic characteristic of BS, especially in the current Covid-19 pandemic era.

## Introduction

Bloom syndrome (BS, MIM 210900) is a rare congenital disorder first described as “congenital telangiectatic erythema resembling lupus erythematosus”^1^ by the dermatologist David Bloom in 1954^2^. Clinical characteristics of the syndrome are primary microcephaly, growth deficiency, short stature, photosensitivity, cancer predisposition, and immunodeficiency^3^. BS is inherited in an autosomal recessive fashion^4^ and is mainly caused by biallelic loss-of-function (LoF) mutations in the *BLM* gene^5,6^. *BLM* is located on chromosome 15q26.1 and encodes one of the five human RecQ helicases, the BLM helicase (RECQL3)^7,8^. The BLM helicase has essential roles during DNA replication and repair processes, which makes it an important component of the cell for the maintenance of genomic stability^9^. Accordingly, cells from BS patients show high levels of genomic instability, e.g., chromosome aberrations and mitotic defects, which overall contribute to the pathogenesis of the disease^2,10^. As an evidence of genomic instability, BS patient cells show elevated rates of sister chromatid exchange (SCE)^11^, which was used as a cytogenetics test in the diagnosis of BS when the molecular diagnostic tools were limited.

The BLM helicase forms part of a four-subunit protein complex, known as the BTR complex or the BLM dissolvasome, which additionally includes the topoisomerase III alpha (TOP3A, MIM 601243) and the RecQ-mediated genome instability protein 1 (RMI1, MIM 610404) and 2 (RMI2, MIM 612426)^12–14^. The BTR complex dissolves DNA intermediates such as G-quadruplexes, D-loops, and Holliday junctions, which occur during different steps of the homologous recombination repair pathway^8,15,16^. Furthermore, the BTR complex is the only known protein complex that can dissolve Holliday junctions without the occurrence of crossovers between the homologous DNA strands^17,18^. Biallelic LoF mutations, small and large indels, as well as mutations affecting correct splicing of *BLM* were described in BS patients^2,19^. Moreover, LoF mutations in other members of the BTR complex, i.e., *TOP3A, RMI1*, and *RMI2*, have been recently linked to BS^6,20,21^. The phenotypic characteristics of patients with biallelic *TOP3A* mutations were similar to those of patients with recessive *BLM* mutations in terms of primary microcephaly, growth retardation, and café-au-lait spots^4,20^. However, dilated cardiomyopathy, which is not a typical characteristic of BS, was also reported in *TOP3A*-mutated patients^20^. Another study reported a large homozygous deletion covering the whole *RMI2* gene to be associated with a BS phenotype in two patients and the diagnosis was confirmed by increased SCE rates although the phenotypic features of the patients were milder than in the common BS phenotype^21^.

Here, we report a patient cohort of eight individuals from six families presenting with main characteristics of BS phenotype. We identified one novel and two previously described homozygous, deleterious mutations in *BLM* in six individuals and defined the clinical phenotype of patients with a homozygous LoF variant in *RMI1* that has recently been reported^20^. We describe the molecular and clinical findings of the patients and compare the phenotypic expression.

## Materials and Methods

### Subjects

All subjects or their legal representatives gave written informed consent to the molecular genetic analyses and the publication of the results. This study was performed according to the Declaration of Helsinki protocol and approved by the local institutional review board (University Medical Center Göttingen, Germany). Samples of genomic DNA from participating family members were extracted from peripheral blood lymphocytes in EDTA solution by standard extraction procedures. In the physical exam of the patients, the height, weight, and occipitofrontal head circumference (OFC) measurements were recorded and evaluated according to the percentile values of the corresponding age groups in Turkish children^22^.

### *BLM* mutation screening

Genomic DNA samples of three patients were screened directly for mutations in *BLM* (NM_000057.4) using specific primers for each exon of the gene including the exon-intron boundaries. Following amplification, Sanger sequencing of the amplified regions was done by BigDye Terminator v3.1 (Applied Biosystems) method on an ABI 3500XL sequencer and the results were visualized using the 4Peaks software^23^. When a mutation was detected, the parents were also investigated for the corresponding region in the same way.

### Microcephaly panel sequencing

An NGS-based in-house microcephaly panel (Agilent Technologies, Santa Clara, CA) covering 78 microcephaly-related genes including the intron-exon boundaries was used for the diagnosis of one patient (www.humangenetik-umg.de). DNA was extracted from blood and enriched with the SureSelect QXT method, PCR-amplified, and sequenced on the Illumina NextSeq system. The sequencing data was evaluated in comparison to the reference sequence (from www.ensembl.org) with the Sequence Pilot software (jsi medical systems GmbH; Version 5.0.0 Build 506). Detected unknown variants were evaluated by computer programs such as SIFT^24^ (https://sift.bii.a-star.edu.sg), Mutationtaster^25^ (www.mutationtaster.org), PolyPhen2^26^ (http://genetics.bwh.harvard.edu/pph2/), M-CAP^27^ (http://bejerano.stanford.edu/mcap/), Human Splicing Finder (http://www.umd.be/HSF3/HSF.shtml), BDGP Splice Site Prediction^28^ (https://fruitfly.org/seq_tools/splice.html), or with the Alamut program (Interactive Biosoftware). The identified BS phenotype-related variants were confirmed via Sanger sequencing as explained above.

### Whole-exome sequencing

Whole-exome sequencing (WES) was performed on four individuals from three different families. The DNA samples from blood were enriched using Agilent SureSelect Human All ExonV6 r2 kit and sequenced on an Illumina HiSeq4000 sequencer. The variants were analyzed using the “Varbank” pipeline from Cologne Center for Genomics (CCG) by applying the following filter criteria^29^: coverage of >6 reads, quality score of >10, allele frequency ≥25%, and a minor allele frequency <0.1% in the 1000Genomes database and the Exome Variant Server (EVS; NHLBI Exome Sequencing Project). The resulting variants were considered as candidates when the phenotype-genotype link was built.

### Variant confirmation by Sanger sequencing

All variants detected by NGS-based techniques were confirmed by the standard Sanger sequencing method as explained above.

## Results

In this study, we assessed the clinical phenotypes and analyzed the molecular basis of eight patients with Bloom syndrome from six families.

### Mutational spectrum of Bloom syndrome patients

The molecular analyses revealed three distinct homozygous mutations in *BLM* in six patients from five families (Table 1). Additionally, a 5-bp deletion in *RMI1* was detected in two cousins from a consanguineous family^20^. All families originated from same country and consanguinity between parents was reported in three families with homozygous *BLM* mutation patients. In one patient, F1/II-4, we identified by WES a novel homozygous mutation in *BLM* (c.572_573delGA; p.Arg191Lysfs*4) and confirmed this causative variant by Sanger sequencing. In this consanguineous family, the parents were heterozygous carriers (Fig. 1a). The prediction of this variant on transcription level was the introduction of a premature stop codon in the third exon resulting in a truncated protein that would lack important domains of the helicase enzyme, e.g., ATP-binding domain and the helicase domain^30^ (Fig. 1b and 1c). This novel mutation was also identified in another patient, F2/II-2, in homozygous state by using an in-house built microcephaly multigene-panel test. The results were confirmed by Sanger sequencing and the parents were heterozygous for the mutation.

**Table 1.**
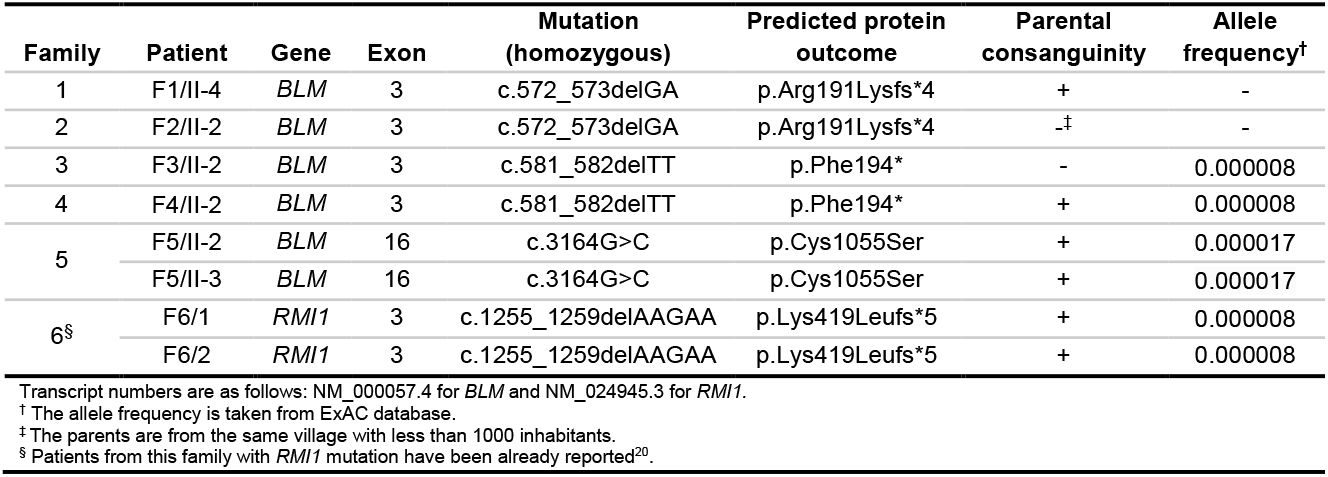
Mutations identified in *BLM* and *RMI1* genes in patients with Bloom syndrome phenotype.

**Figure 1.**
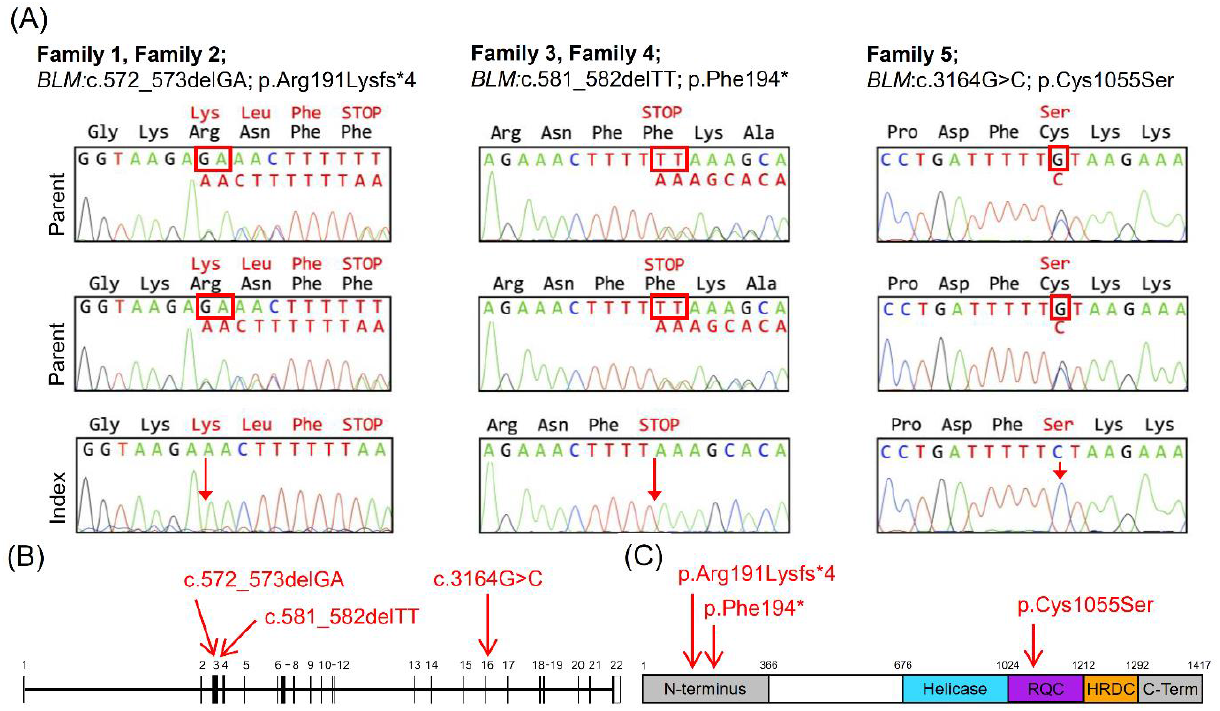
Molecular characterization of detected *BLM* mutations. **(a)** Chromatograms of *BLM* mutations from gDNA samples of representative family members, e.g., parents and index show the heterozygous and homozygous state for each disease-causing mutation. The parents of all index patients were heterozygous carriers and all the patients were homozygous for the detected mutations in *BLM*. **(b)** Positions of the identified mutations are shown in reference to the schematic representation of *BLM* (NM_000057) shown with exons (dark boxes), introns (vertical lines), and UTRs (open boxes). **(c)** Conserved domains of BLM protein are shown with colors as follows: blue, helicase domain; purple, ion (Zn^2+^) binding site and RecQ C-terminal domain; and orange, ribonuclease D C-terminal domain. The C- and N-terminal ends of the protein are shown in grey color.

A known homozygous nonsense mutation in *BLM* (c.581_582delTT;p.Phe194*)^31^ was identified in patient F3/II-2 by WES analysis. In the ExAC database, the allele frequency of this variant was 8 × 10^−6^ (rs367543026) and was predicted to be pathogenic. The same variant was detected in another patient (F4/II-2), in whom the *BLM* coding regions were directly sequenced as the clinical diagnosis of BS had already been given. Furthermore, molecular investigation of *BLM* revealed a homozygous missense variant (c.3164G>C;p.Cys1055Ser)^5^ in two siblings (F5-II/2 and F5-II/3) whose consanguine parents were heterozygous carriers. This missense mutation was located in exon 16 and had an allele frequency of 17 × 10^−6^ (rs367543029). The position of the altered cysteine was on the C-terminal part of the well-conserved RecQ domain, which is important for the ion binding and the helicase enzyme activity of the BLM protein^32^ (Fig. 1c).

Additionally, by performing WES in a *BLM* mutation-negative family with BS, we detected a homozygous truncating variant in *RMI1* in two siblings presenting a BS phenotype and included their phenotypic examination into the present study^20^. The parents of both siblings were related as step-cousins and were heterozygous carriers for the detected variant. No other variant was detected which could explain the disease phenotype.

### Clinical findings in Bloom syndrome patients

We reinvestigated the clinical features of our patients in order to determine the spectrum of *BLM*- and *RMI1*-associated phenotypes. All the index patients included in this study suffered from intrauterine growth deficiency and had a low fetal head circumference (Table 2). Some of the prominent features of BS such as microcephaly and growth deficiency could be already observed in the prenatal period. After birth, which mostly occurred at term, microcephaly and growth retardation continued in all patients resulting in a global developmental delay and short stature, which was in line with the current clinical synopsis of BS^6^ (Table 3). All patients were examined in either childhood or early adolescence and were mainly referred to our clinic due to growth retardation. The most severe case of growth deficiency and microcephaly was patient F4/II-2, presenting with -6 SD in height, -4.9 SD in weight, and -7.6 SD in head circumference. All other patients shared a similar pattern of postnatal growth failure for all three parameters (Table 3). In addition, patient F1/II-4 received growth hormone treatment for three years in early childhood, however, the growth values stayed low.

**Table 2.**
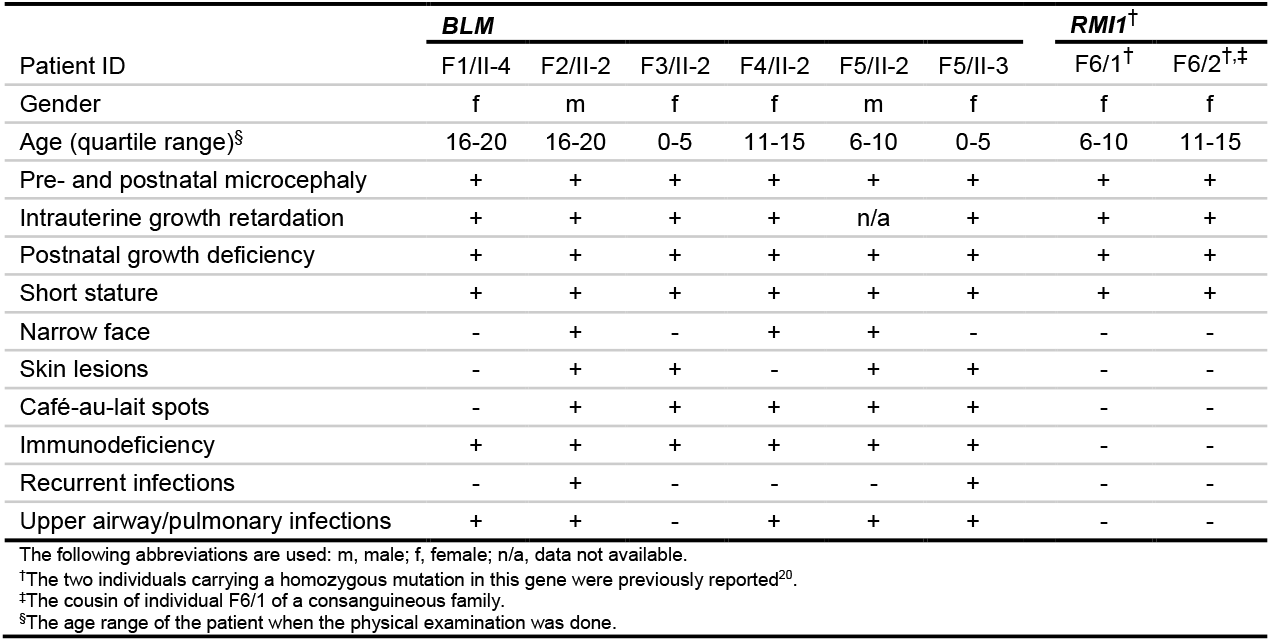
Clinical findings of patients with homozygous mutations in *BLM* and *RMI1* genes.

**Table 3.** Details of clinical findings of patients with Bloom syndrome phenotype.

| Mutated Gene | Patient ID | Gender | Growth Parameters |  |  |  |  |  |  |  | Immunological Parameters |  |  |  |
| --- | --- | --- | --- | --- | --- | --- | --- | --- | --- | --- | --- | --- | --- | --- |
|  |  |  | Birth |  |  |  | Postnatal |  |  |  | IgG | IgA | IgM | IgG1 |
|  |  |  | Term of birth | Length z-score (SD) | Weight z-score (SD) | OFC z-score (SD) | Age at Exam (quartile range) | Height z-score (SD) | Weight z-score (SD) | OFC z-score (SD) |  |  |  |  |
| *BLM* | F1/II-4<sup>†</sup> | female | at term | -4.3 | -5 | n/a | 16-20 | -3,9 | -5,5 | -6 | N | low | low | n/a |
| *BLM* | F2/II-2 | male | 30 weeks | n/a | N | n/a | 16-20 | -4.6 | -4,7 | -5,1 | low | low | low | N |
| *BLM* | F3/II-2 | female | n/a | n/a | n/a | n/a | 0-5 | -4,2 | -3,2 | -4,7 | low | low | low | low |
| *BLM* | F4/II-2 | female | at term | N | -3,3 | n/a | 11-15 | -6 | -4,9 | -7,6 | low | n/a | n/a | n/a |
| *BLM* | F5/II-2<sup>‡</sup> | male | at term | N | -2,7 | n/a | 6-10 | -1,8 | -3,6 | -3,1 | n/a | low | low | n/a |
| *BLM* | F5/II-3 | female | 10 days late | N | N | -3,2 | 0-5 | -2,3 | -4,2 | -5,1 | low | low | low | N |
| *RMI1* | F6/1<sup>§</sup> | female | 34 weeks | n/a | -1,7 | n/a | 6-10 | -4 | -3.8 | -5 | n/a | n/a | n/a | n/a |
| *RMI1* | F6/2<sup>§</sup> | female | 38 weeks | n/a | -3,1 | n/a | 11-15 | -2,9 | -0,9 | -5 | n/a | n/a | n/a | n/a |

The z-scores are calculated using the Turkish children's mean for age and gender. The following abbreviations are used: n/a, data not available; N, normal.
<sup>†</sup>The individual received growth hormone treatment in early childhood.
<sup>‡</sup>Wilms tumor was detected and operated in early childhood.
<sup>§</sup>These individuals have been already reported<sup>20</sup>.

A narrow face, sunlight sensitivity, and café-au-lait spots were observed on some but not all of the patients with homozygous *BLM* mutations. Café-au-lait spots in the form of hypo- and hyperpigmentation were common among the *BLM*-mutated patients except for one patient. On the other hand, sunlight-sensitive skin lesions, which are considered as a major phenotypic characteristic of BS, were present only in two of our *BLM*-mutated patients (F2/II-2 and F3/II-2). Interestingly, neither of our two patients with *RMI1* mutation showed any dermatological characteristics of BS.

### Immunodeficiency is prominent in patients with *BLM* mutations

Immunodeficiency by means of low immunoglobulin levels (e.g., IgG, IgA, IgM, and/or IgG1) was detected in all patients carrying homozygous mutations in *BLM* (Table 3). Accordingly, recurrent infections were an important finding for the clinical synopsis of BS patients. These infections occurred mostly on the upper airway tract. Some patients, e.g., F1/II-4, F4/II-2, and F5/II-2, suffered from more severe repetitive pulmonary infections. The most severe cases of chronic lung infections were patient F5/II-2 at childhood age who experienced bronchitis every year, and patient F1/II-4, who suffered from influenza 3-4 times per year until adolescence. In contrast, both *RMI1*-associated BS patients were not reported to have immunodeficiency and neither of them experienced recurrent infections.

### Bloom syndrome phenotype varies among patients with different mutations

In line with the clinical definition of BS, which includes primary microcephaly and growth delay, all of our eight patients presented pre-/postnatal microcephaly and short stature^2^. However, dermatological features, e.g., photosensitivity and café-au-lait spots, were heterogeneous characteristics among the *BLM*-mutated patients. Of note, neither of the patients with *RMI1* mutation showed any skin phenotype of BS. Similarly, immunodeficiency was presented by the patients with homozygous *BLM* mutations but not by *RMI1*-mutated patients. Overall, we highlight that among our patient cohort, the individuals with homozygous mutations in *BLM* showed more severe BS phenotypic features than the individuals with the homozygous *RMI1* mutation.

## Discussion

Evaluation of phenotypes and mutational spectra of rare diseases such as Bloom syndrome often faces the challenge of a small number of patients. Although the disease was described in 1954^1^, the most comprehensive BS patient cohort was reported in 2007^6^. The study reported 134 BS patients and extensively described the causative mutations in *BLM*^6^. Yet, 9 out of 134 BS patients lacked a disease-causing mutation in *BLM*, indicating a likely genetic heterogeneity^2,6^. In this study, we report a patient cohort of eight individuals diagnosed with BS. The molecular analyses revealed one novel and two previously described LoF mutations in *BLM* in homozygous state in six of the patients. Moreover, we included a previously described homozygous truncating variant in *RMI1* into our study^20^. Our molecular diagnosis algorithm specific for BS comprises *BLM* gene screening, panel screening, and/or WES. The causative mutations that were detected via NGS-based technique were confirmed by Sanger sequencing in the patients and the parents. This methodology, which includes NGS-based approaches, could be considered as a new standard for the molecular diagnosis of Bloom syndrome since other genes than *BLM* are associated with BS phenotype, e.g., *TOP3A, RMI1*, and *RMI2*^20,21^. Therefore, further associations of the BS phenotype to genes involved in various pathways of maintenance of genomic stability might be expected and WES analysis would thus be of high interest to identify such novel links. As an example, we hypothesize that patients in the Bloom syndrome registry^6^ with no detected *BLM* mutations likely have one or more causative mutations in other genes or have at least intronic deleterious variants in *BLM*, which could be detected by next-generation sequencing techniques.

In the current study, one novel frameshift mutation in *BLM* (c.572_573delGA;p.Arg191Lysfs*4) was detected by WES analysis in one BS patient. The variant was identified as damaging by various in-silico prediction tools due to a premature stop codon in the N-terminal domain of the protein. Therefore, we report this homozygous variant as a disease-causing mutation in *BLM* detected in two individuals for the first time. Although the two patients were from different families, we hypothesize a founder effect for this variant, considering the high rates of consanguinity in the country of origin. Parents of Family 1 reported consanguinity from second degree. Parents of Family 2 claimed no relation although both were originating from the same village with less than 1,000 inhabitants. Therefore, we assume a common ancestor for this variant. Furthermore, a known mutation (*BLM*: c.581_582delTT;p.Phe194*) was detected in homozygous state in two BS patients coming from unrelated families. In an earlier study, this mutation was reported homozygously in one individual with BS phenotype^31^. The 2-bp deletion was deleterious due to the introduction of a premature stop codon, causing a truncated protein with missing enzymatic domains (Fig. 1c). The third mutation detected in *BLM* was the extensively studied (c.3164G>C;p.Cys1055Ser) missense mutation^5^. This mutation affects one of the four highly-conserved cysteine domains which bind zinc ion^32,33^. The ion binding to this domain is essential for the ATPase and helicase enzyme activity as well as for the stabilization of the BLM protein^34,35^. Therefore, this amino acid change was detrimental to the enzyme activity and considered to be deleterious as a variant, resulting in the BS phenotype.

Cancer predisposition is an important clinical feature in BS as it greatly affects patients’ life span and quality. In our patient cohort, we have not detected any malignancy except one incident of Wilms tumor in one patient who underwent surgery in early childhood. The lack of cancer incidence in our patient cohort could be due to the young age of the patients: our oldest patient was at adolescent ages, while the mean age of onset of cancer in BS patients is 24 years^6^. In addition, patient F1/II-4 received growth hormone treatment for three years; however, the growth parameters did not change. The BS characteristics of this patient were heterogeneous as skin lesions and a narrow face were missing and, therefore, growth hormone treatment was started before the completion of molecular diagnosis. We want to note that great care must be taken while deciding on growth hormone treatment, bearing in mind the possibility of malignancy increase for BS patients with pronounced cancer predisposition^36^.

We observed a phenotypic variety among the BS patients depending on the causative mutations. The strongest phenotypic variance among the BS patients was observed between patients with homozygous LoF mutations in different genes, namely *BLM* and *RMI1*. The patients with homozygous *BLM* mutations showed a more severe BS phenotype in terms of photosensitivity and immunodeficiency in comparison to the *RMI1*-mutated patients. The gene products of the two genes; BLM helicase and RMI1 proteins, form part of a protein complex named the BTR complex, which dissolves DNA intermediates. Once the complex is near the chromatin, BLM helicase acts on the branches of entangled DNA by moving them closer to form a hemicatenated structure^17,37^. Then, the topoisomerase III alpha protein separates the two strands by nicking to generate the final non-crossover products^38^. RMI1 stimulates the process by associating with both BLM and TOP3A^12,39^. The major role within the complex may possibly be assigned to BLM and TOP3A proteins since they are the enzymatic parts acting on the DNA^40^. Nevertheless, lack of RMI1 protein has been shown to lead to defective cell proliferation due to deficient phosphorylation of BLM^13,41^. In summary, we conclude that mutations in the different members of the BTR complex have a varying impact on the severity of the BS phenotype, especially when comparing *BLM* and *RMI1-*associated phenotypes.

A distinct phenotypic characteristic of BS is the pronounced immunodeficiency profile^42^. Strikingly, all our patients with homozygous *BLM* mutations showed immunity problems resulting in recurrent infections mainly of the upper respiratory tract. However, patients with the homozygous *RMI1* mutation did not show this clinical feature. Furthermore, we examined two siblings with BS in one family and observed the phenotypic variance based on a sibship. Infection rates were higher for the younger patient, although both children suffered from immunodeficiency and had similar BS characteristics. Based on our evaluations, we emphasize that the frequent infections in BS are as important as any other life-threatening element of BS like cancer predisposition. Due to the low levels of immunity, BS patients should be considered susceptible to any infection such as the Covid-19 pandemic that arose in 2020.

In summary, in eight patients presented with BS from six families, we detected one novel homozygous frameshift mutation in *BLM* in two patients from two families, two already known mutations in three families, and one homozygous truncating mutation in the *RMI1* gene in a consanguineous family^20^. All variants were homozygous loss-of-function mutations associated with the BS phenotype. Moreover, patients with homozygous mutations in the *BLM* gene showed more severe BS phenotype characteristics in terms of photosensitivity, immunodeficiency, and infection rate than the patients carrying the homozygous *RMI1* variant. The immunodeficiency profile of BS is an important characteristic of the disease. Thus, we highlight that extra care must be taken to protect BS patients, especially in the current Covid-19 pandemic.

## Data Availability

The whole-exome sequencing raw data are not publicly available due to privacy or ethical restrictions. Processed genetic data generated or analyzed within this study are available upon request.

## Acknowledgments

We are grateful to all patients for participating in this study and Karin Boss for critically reading the manuscript. This work was supported by the Deutsche Forschungsgemeinschaft (DFG, German Research Foundation) under Research Group FOR 2800 **“**Chromosome Instability: Cross-talk of DNA replication stress and mitotic dysfunction”, SP5 and SPZ to B. Wollnik and Germany’s Excellence Strategy, Cluster of Excellence “Multiscale Bioimaging: from Molecular Machines to Networks of Excitable Cells” (MBExC; EXC 2067/1-390729940) to B. Wollnik.

## Data Availability Statement

Pedigrees and family histories are available upon request. The whole-exome sequencing raw data are not publicly available due to privacy or ethical restrictions. Processed genetic data generated or analyzed within this study are available upon request.

## Ethical Statement

Written informed consent of all participants or their legal representatives was obtained prior to participation in the study. This study was performed according to the Declaration of Helsinki protocol and approved by the local institutional review board (University Medical Center Göttingen, Germany) under approval number 3/2/16.

